# Diagnostic Performance of multimodal biomarker in colorectal cancer

**DOI:** 10.1101/2025.08.01.25332760

**Authors:** Shuang Yang, Yu Qing Wang, Jiang Li, Kai Wu Xu

## Abstract

**Objective:** This study aimed to evaluate the diagnostic performance of plasma methylated SEPT9 (mSEPT9) gene combined with carcinoembryonic antigen (CEA), carbohydrate antigen 199 (CA199), fecal occult blood test (FOBT), red blood cell distribution width (RDW), and inflammation-related indices from complete blood count (CBC) for colorectal cancer (CRC).

**Methods:** A prospective study was conducted on 188 patients pathologically diagnosed with CRC (CRC group) and 693 control subjects with gastrointestinal symptoms but non-CRC diagnoses (control group) admitted to Hunan Provincial People’s Hospital from January 01, 2024 to December 31, 2024. Data on mSEPT9, CEA, CA199, FOBT, and CBC were collected. Binary logistic regression analysis was used to establish a predictive model for CRC risk factors, and receiver operating characteristic (ROC) curve analysis was performed to evaluate the diagnostic performance of each index.

**Results:** The positive rates of mSEPT9 and FOBT in the CRC group were significantly higher than those in the non-CRC group (*P<*0.001). The levels of CEA, CA199, RDW-CV, RDW-SD, neutrophil-to-lymphocyte ratio (NLR), and platelet-to-lymphocyte ratio (PLR) were significantly higher in the CRC group than in the control group (*P<*0.001), while the lymphocyte-to-monocyte ratio (LMR) was significantly lower (*P<*0.001). Univariate logistic regression analysis showed that mSEPT9, CEA, CA199, FOBT, RDW-CV, RDW-SD, NLR, and PLR were independent predictive risk factors for CRC. Multivariate regression analysis indicated that patients with positive or elevated mSEPT9, CEA, CA199, FOBT, RDW-CV, and PLR were more likely to have CRC. The combined model of mSEPT9, CEA, CA199, FOBT, RDW-CV, and PLR demonstrated an impressive area under the ROC curve (AUC) of 0.939, with a sensitivity of 0.920 and specificity of 0.839, highlighting excellent screening efficacy for CRC.

**Conclusion:** A screening model incorporating mSEPT9, CEA, CA199, FOBT, RDW-CV, and PLR provides valuable insights for CRC diagnosis.

## Introduction

Colorectal cancer (CRC) is a prevalent gastrointestinal malignant tumor globally, posing a severe threat to human health and exhibiting an increasing trend of younger onset^[1]^. The statistical data on Chinese malignant tumors released by the China Cancer Center in 2024 shows that the incidence rate of CRC has ranked second among all malignant tumors in China, with the mortality rate ranking fourth^[2]^. Early-stage CRC has a relatively high treatment success rate, but CRC patients rarely present with corresponding symptoms in the early stage. Most CRC patients are diagnosed at an advanced stage, leading to poor prognosis. Therefore, early screening of CRC is a key factor in improving clinical treatment efficacy and reducing CRC mortality ^[3-4]^. Common methods for CRC screening and diagnosis include fecal occult blood test (FOBT) based on immunochemical analysis, serum tumor markers, colonoscopy, etc. ^[5]^. However, the sensitivity or specificity of single FOBT or serum tumor marker testing is still unsatisfactory, while colonoscopy is invasive, requires complex intestinal preparation, and may be associated with risks such as bleeding and perforation, resulting in low compliance. Therefore, there is an urgent clinical need to develop a non-invasive, highly sensitive, and specific screening protocol for the early diagnosis of CRC^[6]^, especially for individuals who are reluctant to undergo colonoscopy.

DNA methylation, one of the most common epigenetic modifications, plays a pivotal role in the occurrence and development of various cancers, including CRC^[7]^. A prospective cohort study has validated the effectiveness of DNA methylation-based blood tests for CRC screening in high-risk populations, demonstrating that such tests even outperform traditional markers like CEA and CA199^[8]^. Currently, approved methylation-based diagnostic biomarkers for CRC include SEPT9, NDRG4, BMP3, etc.^[9]^. Among these genes, SEPT9 is widely present in eukaryotic cells. The SEPT9 protein encoded by this gene participates in biological processes such as cell division, vesicle transport, and cell membrane remodeling, influencing tumorigenesis. As a tumor suppressor gene, its methylation leads to abnormalities in cell division, thereby contributing to cancer development^[10]^. Studies have shown that overexpression of mSeptin9 can be observed in CRC tissues^[11]^, and it is also detectable in blood samples of CRC patients. Its levels are closely associated with the progression of multiple malignant tumors, including CRC and gastric cancer^[12]^, offering new insights for non-invasive diagnosis of CRC.

In recent years, accumulating studies have revealed a close association between tumorigenesis/progression and inflammatory responses. Inflammation can alter the tumor microenvironment (TME) through multiple mechanisms, influencing all stages of carcinogenesis, including initiation, promotion, and progression^[13]^. Inflammation-related indices, such as the neutrophil-to-lymphocyte ratio (NLR), platelet-to-lymphocyte ratio (PLR), and lymphocyte-to-monocyte ratio (LMR), have been demonstrated as prognostic markers for gastrointestinal tumors^[14]^. Meanwhile, red cell distribution width (RDW), a key indicator of erythrocyte volume variability, includes two core parameters: red blood cell distribution width-coefficient of variation (RDW-CV) and red blood cell distribution width-standard deviation (RDW-SD). Studies have shown that this indicator exhibits significant differences between colorectal cancer (CRC) patients and colorectal adenoma patients^[15]^. These results obtained from complete blood count analysis, when combined with mSEPT9 detection, may also enhance the diagnostic efficacy for CRC^[16]^.

Carcinoembryonic antigen (CEA) and carbohydrate antigen 199 (CA199), two serum tumor markers combined with FOBT, are currently common non - invasive examination methods for clinical diagnosis of CRC, but there are still obvious limitations in their diagnostic application for CRC^[17]^. Studies have shown that the combined detection of mSEPT9 with CEA, CA199, NLR and PLR^[18]^ or the combined detection of mSEPT9 with CEA, CA199 and FOBT ^[19]^ can improve the accuracy of CRC diagnosis and reduce the occurrence of missed diagnosis and misdiagnosis. As far as we know, there is no study that has carried out integrative analysis on the predictive and combined diagnostic performance of mSEPT9 with CEA, CA199, FOBT, RDW and CBC inflammation - related indices for CRC. This study intends to construct a binary logistic regression model with these indices to screen the predictive risk factors for CRC occurrence, and use the receiver operating characteristic curve (ROC) to analyze the diagnostic value of these indices for CRC in single and combined detection, so as to provide new strategies for the screening and diagnosis of CRC.

## Materials and Methods

### Study Subjects

A prospective study was conducted by enrolling 188 patients pathologically diagnosed with CRC at Hunan Provincial People’s Hospital from January 01, 2024 to December 31, 2024 as the CRC group, and 693 patients with digestive symptoms (such as abdominal pain, diarrhea, changes in bowel habits, or hematochezia) who underwent relevant treatment and were pathologically diagnosed as non-CRC at the same hospital during the same period as the control group.Inclusion criteria: CRC patients were diagnosed according to the Chinese Colorectal Cancer Diagnosis and Treatment Guidelines (2023 Edition), and complete clinical data were ensured.Exclusion criteria:(1)Female patients who were lactating or pregnant.(2) Patients with other malignant tumors. (3) Patients who had received previous radiotherapy, chemotherapy, immunotherapy, surgical treatment, or systemic therapy.This study protocol was approved by the Ethics Committee of the Hunan Provincial People’s Hospital.All the patients also understood and signed the informed consent.

### Research Methods

#### mSEPT9 Detection (PCR Fluorescence Probe Method)

The detection reagents were purchased from BioChain Institute, Inc., and the detection process was performed according to the kit instructions. (1) Sample collection and storage: Venous blood (10 mL) was collected using an EDTA-K2 anticoagulant vacuum blood collection tube, centrifuged at 1500 r/min for 12 min at room temperature (25°C), and at least 3.5 mL of plasma was separated and stored at -25 to -15°C for detection within 1 week(. 2)DNA extraction and bisulfite conversion from plasma: 3.5 mL of plasma was lysed with lysis buffer, bound to magnetic beads, washed, and eluted to obtain free DNA. Free DNA was subjected to bisulfite conversion, bound to magnetic beads again, washed, dried, and eluted to obtain bisulfite-converted DNA samples (BisDNA)(. 3)Preparation of PCR reaction system: Each PCR reaction required 32 μL of PCR reaction solution and 1.6 μL of polymerase. The PCR reaction solution and polymerase were mixed in the specified ratio in a 2.0 mL centrifuge tube, vortexed thoroughly to ensure uniform distribution, and centrifuged to collect the liquid. 30 μL of the pre-mixed PCR solution was added to a 96-well plate, followed by 30 μL of BisDNA into the corresponding wells. The plate was sealed with a film and centrifuged at 1000 r/min for 1 min(. 4)PCR reaction: Using a real-time fluorescent quantitative PCR instrument (Hongshi SLAN96), the PCR program was set according to the instructions, with the human housekeeping gene actin (ACTB) as the internal reference(. 5)Result interpretation: A positive result was defined as Septin9 Ct ≤ 41.0 and ACTB Ct ≤ 32.1; a negative result was defined as no Septin9 Ct or Septin9 Ct > 41.0 with ACTB Ct ≤ 32.1; an invalid result was defined when ACTB Ct > 32.1 regardless of Septin9 Ct value.

#### FOBT Detection

Fecal occult blood test (FOBT) was performed within 24 hours after stool specimen collection using the colloidal immunochromatographic assay. The detection reagents were purchased from W.H.P.M. Bioresearch & Technology Co., Ltd.

#### CEA and CA199 Detection

Venous blood (3–5 mL) was collected using a vacuum blood collection tube with a separating gel and coagulant accelerator. The sample was centrifuged at 1500 r/min for 12 minutes. The upper-layer serum was transferred to a 2-mL centrifuge tube and stored at -80°C until testing. Detection was performed by chemiluminescence assay, using reagents purchased from Abbott Ireland Diagnostics Division. In this study, the normal reference ranges for CEA and CA199 were 0–5 ng/mL and 0–37 U/mL, respectively. An abnormal result was defined as CEA ≥5 ng/mL or CA199 ≥37 U/mL.

#### Complete Blood Count (CBC) Detection

Fasting venous blood (2 mL) was collected from patients in the morning. Using a SYSMEX-XN9000 hematology analyzer, parameters including RDW-CV, RDW-SD, neutrophil count, platelet count, lymphocyte count, and monocyte count were measured. NLR (neutrophil-to-lymphocyte ratio), PLR (platelet-to-lymphocyte ratio), and LMR (lymphocyte-to-monocyte ratio) were calculated based on the obtained data.These experiments were carried out with the approval of the Ethics Committee of the Hunan Provincial People’s Hospital. All the patients also understood and signed the informed consent.

#### Statistical Analysis

Statistical analyses were performed using SPSS 26.0 software. Measurement data conforming to normal distribution were expressed as mean ± standard deviation (x±s). The t-test was used for comparing measurement data between two groups, while the chi-square test was applied to count data. Non-normally distributed measurement data were presented as median (first quartile, third quartile) [M (Q1, Q3)], and the Mann-Whitney U test was used for intergroup comparisons. Binary logistic regression was employed to analyze factors associated with CRC occurrence. The area under the receiver operating characteristic (ROC) curve (AUC) was used to evaluate the diagnostic efficacy of individual and combined indicators for CRC, and sensitivity, specificity, odds ratios (OR), and 95% confidence intervals (CI) were calculated. A P-value < 0.05 was considered statistically significant.

## Results

### Baseline Clinical Information of the Two Groups

The baseline clinical information of the CRC group and non-CRC control group is shown in Figure 1. A total of 881 participants (564 males and 317 females) were enrolled in this study, including 188 cases in the CRC group (111 males and 77 females, median age 63 years) and 693 cases in the non-CRC control group with similar digestive symptoms (453 males and 240 females, median age 52 years). There was no significant difference in gender composition between the two groups (P>0.05, Figure 1A), while the age difference was statistically significant (*P<*0.001). The CRC group had a higher number of patients aged ≥60 years (Figure 1B).

**Fig 1.**
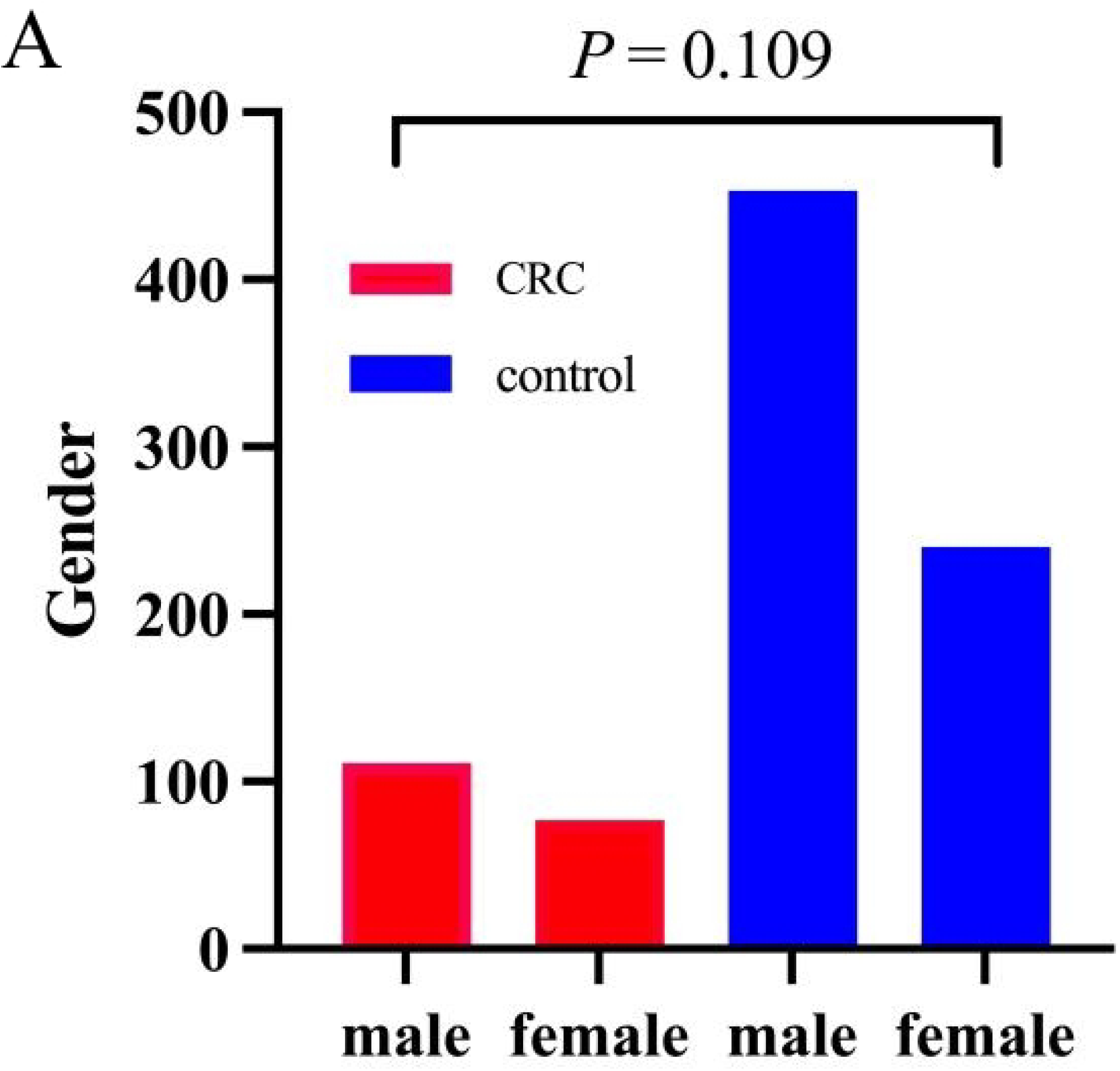
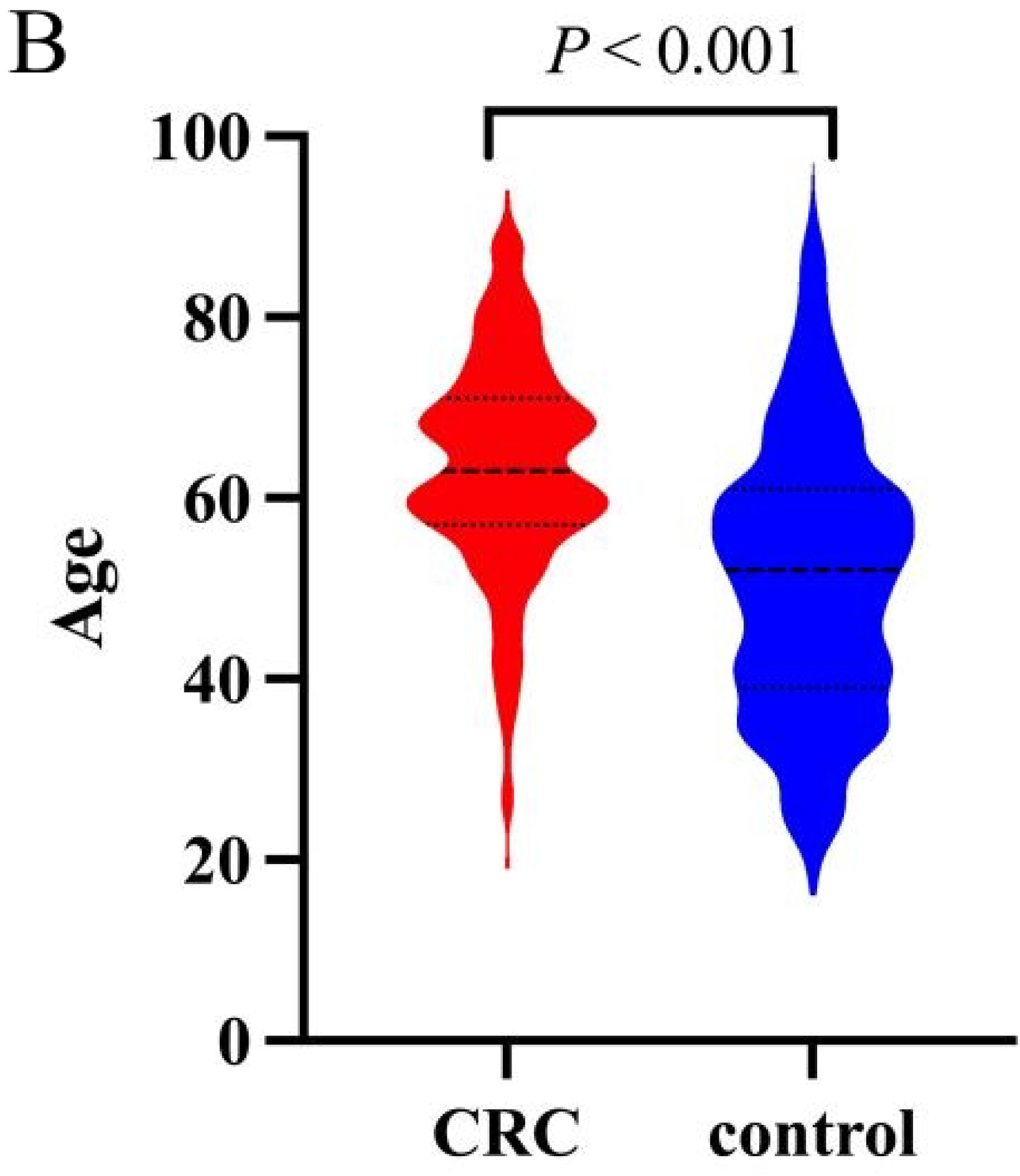
Clinical characteristics of CRC and non-CRC groups. (A)Gender distribution in CRC and non-CRC groups; (B)Age distribution between CRC and non-CRC groups

### Detection Results of mSEPT9 and FOBT in the Two Groups

The positive rates of mSEPT9 and FOBT in the CRC group were 61.2% (115/188) and 85.4% (135/158), respectively, while those in the control group were 5.5% (38/693) and 20.3% (70/344), respectively. The positive rates of both mSEPT9 and FOBT in the CRC group were significantly higher than those in the control group (*P<*0.001, χ^2^ test), as shown in Table 1.

**Table 1.**
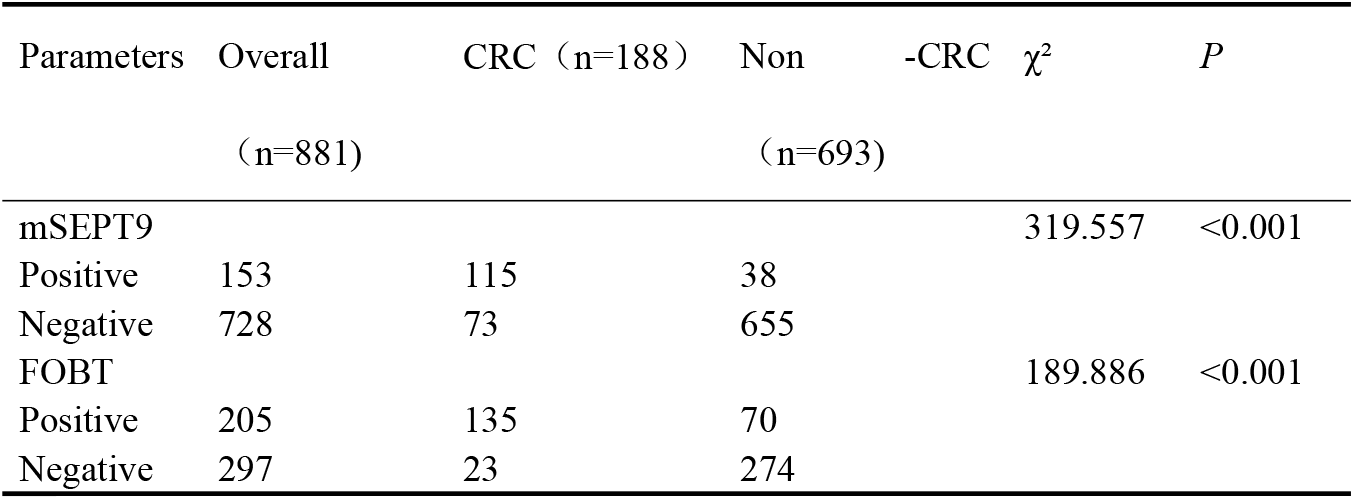
Comparison of mSEPT9 and FOBT between CRC and non-CRC groups.

### Detection Results of CEA, CA199, RDW-CV, RDW-SD, NLR, PLR and LMR in the Two Groups

The seven indicators of CEA, CA199, RDW-CV, RDW-SD, NLR, PLR and LMR in the CRC group and the control group did not all conform to normal distribution, so M (Q1, Q3) was used for statistical description. Mann-Whitney U test was used for analysis. The levels of CEA, CA199, RDW-CV, RDW-SD, NLR and PLR in the CRC group were all higher than those in the control group (*P*<0.001), and the LMR level was lower than that in the control group (*P*<0.001), as shown in Table 2.

**Table 2.** Comparison of CBC and tumor markers between CRC and non-CRC groups.

| Parameters | CRC M (Q1, Q3) | Non -CRC M (Q1, Q3) | Z | P |
| --- | --- | --- | --- | --- |
| CEA,ng/ml | 5.63 (2.50, 15.10) | 1.46 (0.88, 2.22) | -13.819 | <0.001 |
| CA199,U/ml | 13.70 (5.06, 51.35) | 5.93 (3.39, 11.03) | -7.495 | <0.001 |
| RDW-CV | 13.40 (12.70, 14.60) | 12.70 (12.20, 13.40) | -7.683 | <0.001 |
| RDW-SD | 43.80 (40.93, 46.28) | 42.60 (40.30, 45.00) | -3.597 | <0.001 |
| NLR | 3.10 (2.17, 4.95) | 1.99 (1.45, 3.27) | -7.294 | <0.001 |
| PLR | 176.52 (126.80, 245.30) | 129.02 (98.17, 179.36) | -7.175 | <0.001 |
| LMR | 3.26 (2.06, 4.63) | 4.18 (2.75, 5.52) | -4.987 | <0.001 |

### Binary Logistic Regression Analysis for Predicting Risk Factors of CRC Occurrence

Indicators with statistical significance (*P*<0.05) between the CRC group and control group in the above analysis (mSEPT9, FOBT, CEA, CA199, RDW-CV, RDW-SD, NLR, PLR, LMR) were selected as independent variables, and whether suffering from CRC was used as the dependent variable to construct a binary logistic regression prediction model. The results showed that in the univariate binary logistic regression analysis, all 9 above-mentioned indicators were independent predictive factors for CRC occurrence. Among them, 8 indicators including mSEPT9, FOBT, CEA, CA199, RDW-CV, RDW-SD, NLR, and PLR were risk factors for CRC occurrence (OR>1), while LMR was a protective factor for CRC occurrence (OR<1). The above 8 independent risk factors were included in the multivariate binary logistic regression analysis, and the results showed that a total of six indicators, including mSEPT9, FOBT, CEA, CA199, RDW-CV, and PLR, could be used to distinguish whether clinical individuals were more likely to suffer from CRC, as shown in Table 3.

**Table 3.**
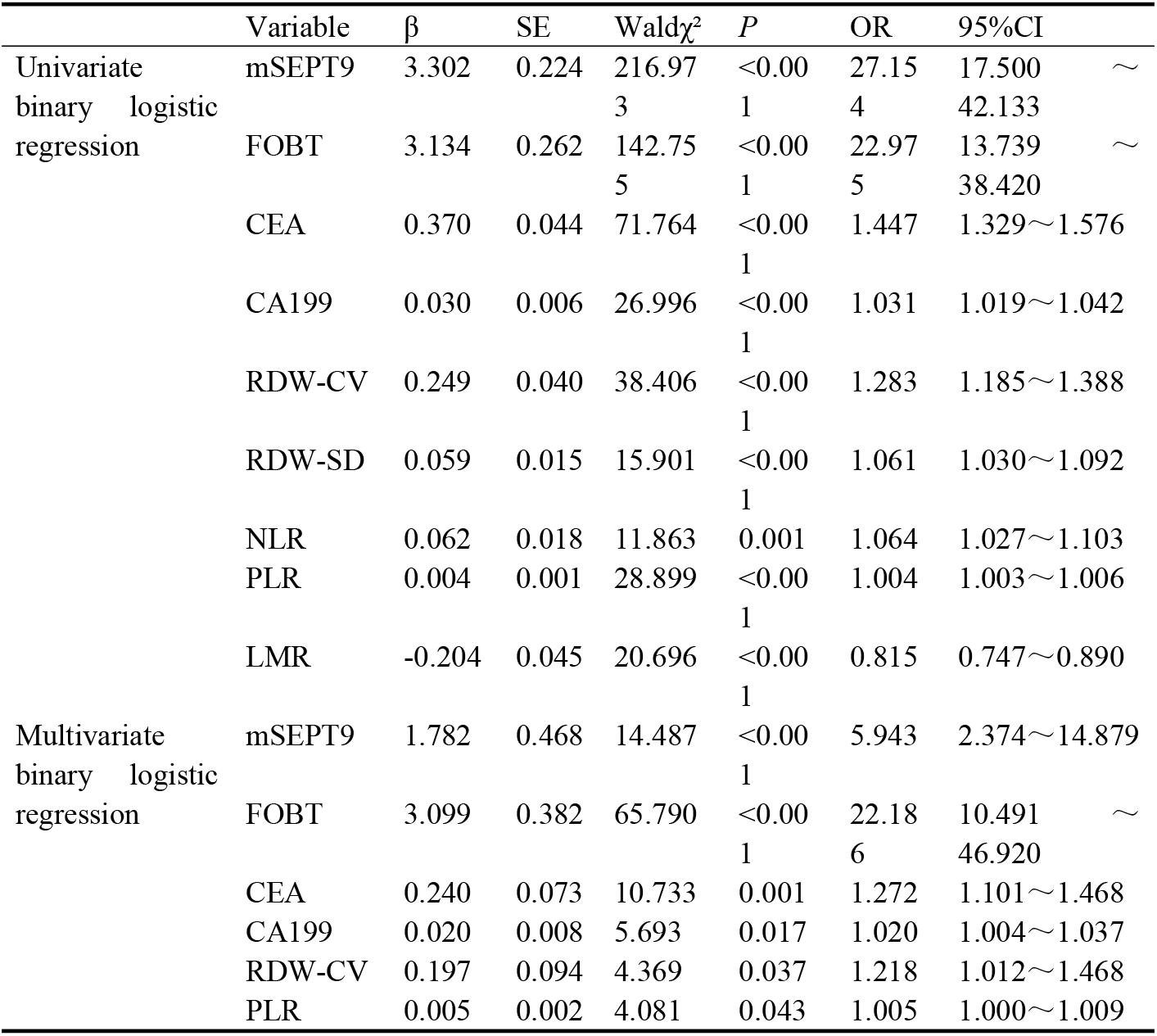
Univariate and multivariate binary logistic regression analysis of CRC risk factors.

**Table 4.**
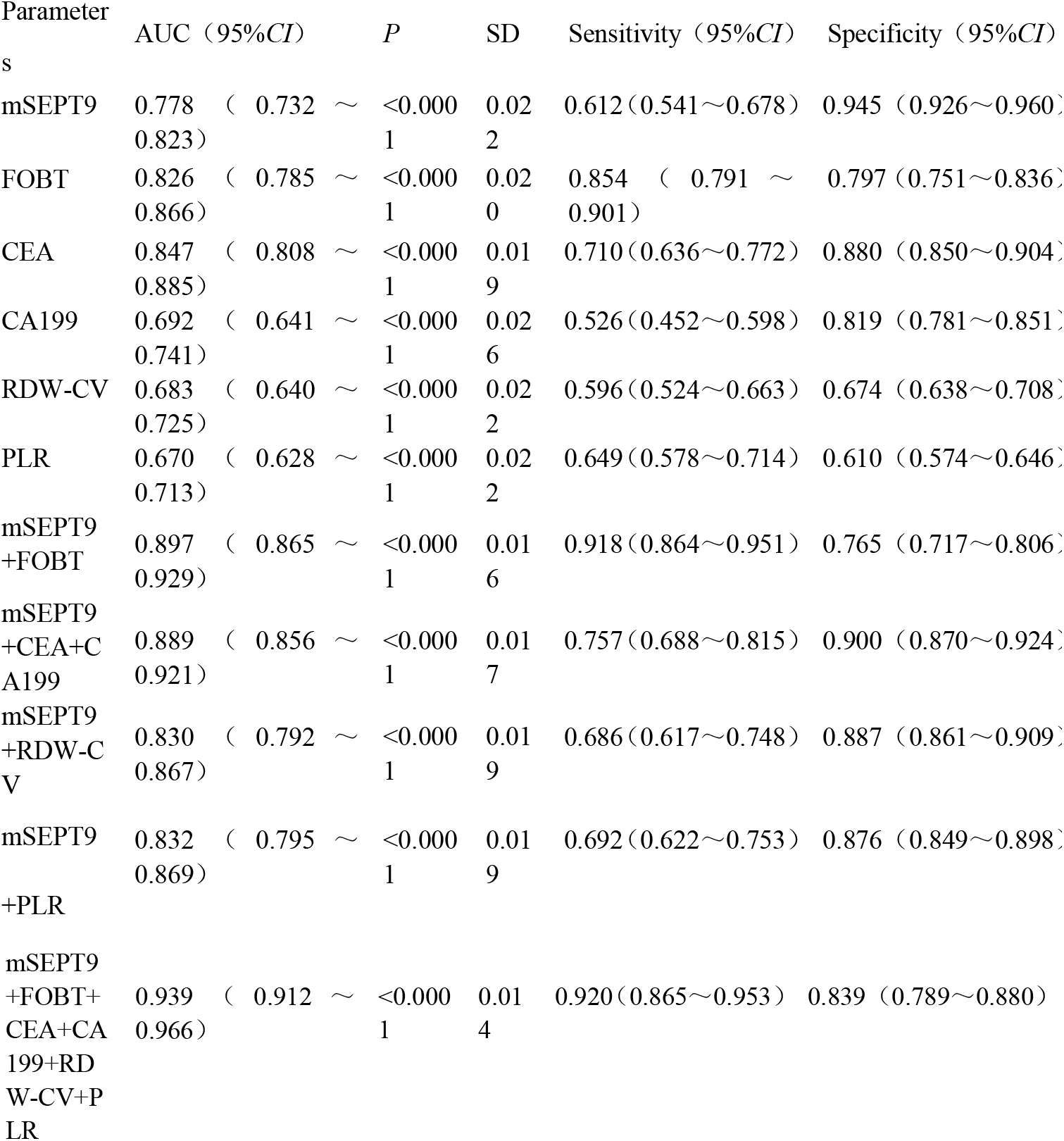
Diagnostic value of mSEPT9 and other biomarkers for CRC detection.

### Diagnostic Efficacy Analysis of mSEPT9, FOBT, CEA, CA199, RDW-CV and PLR for CRC

A diagnostic model was constructed to distinguish between CRC and non-CRC individuals using the predictive risk factors (mSEPT9, FOBT, CEA, CA199, RDW-CV, and PLR) identified by multivariate binary logistic regression analysis. The clinical efficacy of each indicator for independent CRC diagnosis is shown in Table 2. The sensitivities of mSEPT9, FOBT, CEA, CA199, RDW-CV, and PLR were 0.612 (95% CI 0.541–0.678), 0.854 (95% CI 0.791–0.901), 0.710 (95% CI 0.636–0.772), 0.526 (95% CI 0.452–0.598), 0.596 (95% CI 0.524–0.663), and 0.649 (95% CI 0.578–0.714), respectively. The specificities were 0.945 (95% CI 0.926– 0.960), 0.797 (95% CI 0.751–0.836), 0.880 (95% CI 0.850–0.904), 0.819 (95% CI 0.781–0.851), 0.674 (95% CI 0.638–0.708), and 0.610 (95% CI 0.574–0.646), respectively. The AUCs (95% CI) were 0.778 (0.732–0.823), 0.826 (0.785–0.866), 0.847 (0.808–0.885), 0.692 (0.641–0.741), 0.683 (0.640–0.725), and 0.670 (0.628– 0.713), respectively. The ROC curves for independent CRC diagnosis by each indicator are shown in Figure 2A.

**Fig 2.**
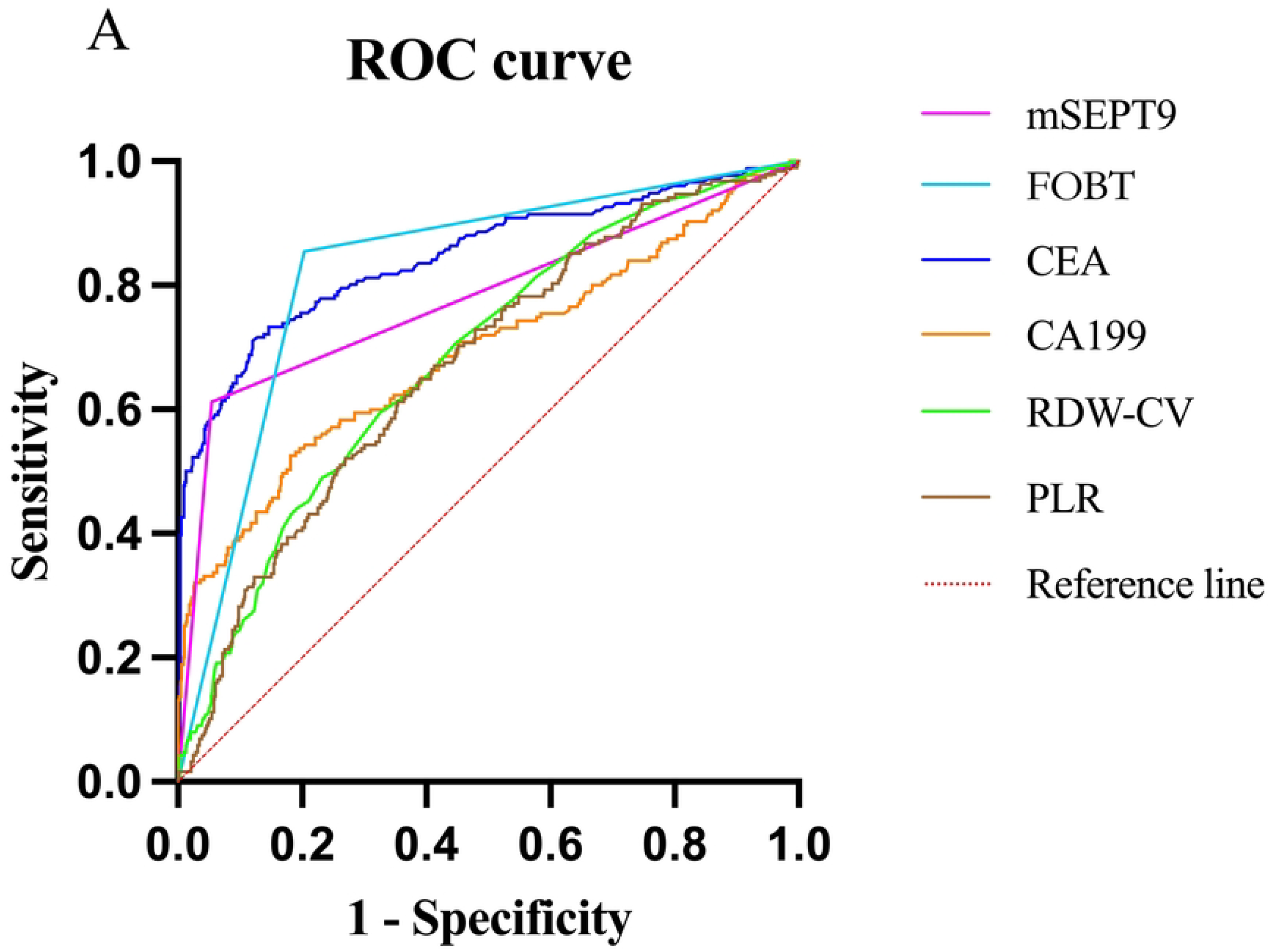
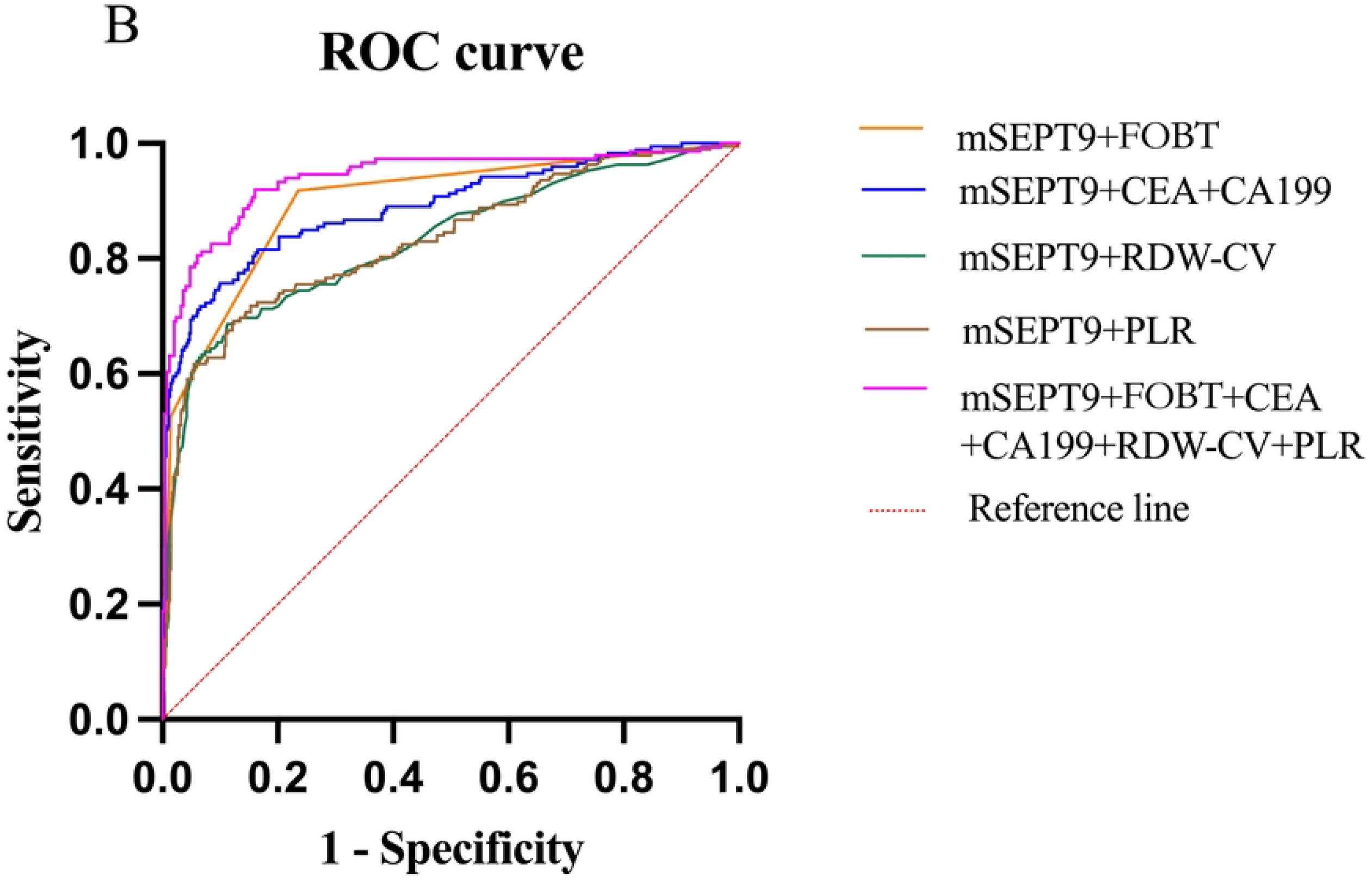
ROC curve analysis of mSEPT9, FOBT, CEA、CA199、RDW-CV、PLR for CRC diagnosis.(A)The diagnostic values of mSEPT9,FOBT, CEA, CA199、RDW-CV、PLR for detecting CRC;(B)The diagnostic values of the combination of mSEPT9, FOBT, CEA, CA199、RDW-CV、PLR for detecting CRC

The diagnostic performance of mSEPT9 combined with other indicators is shown in Table 2. When mSEPT9 was combined with FOBT, the sensitivity increased to 0.918 (95% CI 0.864–0.951), but the specificity decreased to 0.765 (95% CI 0.717– 0.806) compared with individual detections. The combination of mSEPT9 with tumor markers CEA and CA199 improved the sensitivity for CRC diagnosis to 0.757 (95% CI 0.688–0.815), with a slightly lower specificity of 0.900 (0.870–0.924) than that of mSEPT9 alone. The combination of mSEPT9 with RDW-CV and PLR showed similar and relatively low sensitivity and specificity. When mSEPT9 was combined with FOBT, CEA, CA199, RDW-CV, and PLR, the diagnostic efficacy for CRC was the highest, with the AUC (95% CI) increasing to 0.939 (0.912–0.966), and the sensitivity (95% CI) and specificity (95% CI) reaching excellent levels of 0.920 (0.865–0.953) and 0.839 (0.789–0.880), respectively. The ROC curves for combined CRC diagnosis by each indicator are shown in Figure 2B.

## Discussion

Common symptoms of CRC include changes in bowel habits, rectal bleeding, abdominal pain, abdominal distension, anemia, and unexplained weight loss. Most patients are diagnosed at the middle or advanced stages when symptoms appear. In the CRC patient population, early diagnosis and treatment are crucial for improving the five-year survival rate. The CAN-SCREEN (Colorectal cANcer SCReening Economics and adherENce) model indicates that among non-invasive CRC screening strategies, those with higher compliance lead to more favorable health outcomes, manifested as averted CRC deaths, avoided CRC cases, and improved quality of life (LYG)^[20]^. In light of this, selecting a highly compliant and efficient diagnostic method is of utmost importance and urgency.

The Septin9 gene is primarily located at chromosome 17q25.3 and encodes Septin9 protein, a cell cycle-related protein that coordinates motor proteins and myosins during cell division. When the Septin9 gene is silenced by methylation, normal cytokinesis is disrupted, promoting uncontrolled cell proliferation and leading to malignant tumorigenesis^[21]^. mSEPT9 has shown high diagnostic value in multiple malignancies, including liver cancer, gastric cancer, esophageal cancer, lung cancer, and ovarian cancer, with particularly extensive application in CRC diagnosis^[22]^. This technique provides a promising approach for non-invasive assessment and monitoring of colorectal cancer, overcoming multiple limitations of traditional screening methods. In this study, the positive rate of mSEPT9 detection in the CRC group (61.2%) was significantly higher than that in the control group (5.5%). The area under the curve (AUC, 95% CI) of plasma mSEPT9 detection for independent CRC diagnosis was 0.778 (0.732–0.823), indicating moderate diagnostic value, which is consistent with the results of multicenter studies by other researchers ^[23-24]^.

When CRC progresses to a certain extent, rectal bleeding may occur, so fecal occult blood testing (FOBT) using colloidal gold immunodetection remains one of the diagnostic methods for screening average-risk individuals for CRC^[25]^. Fecal immunochemical analysis is inexpensive and more cost-effective than colonoscopy in CRC screening and diagnosis, helping to reduce cancer cases and deaths ^[26-27]^. In this study, the positive rate of FOBT in the CRC group (85.4%) was higher than that in the control group (20.3%). The AUC (95% CI) of FOBT for independent CRC diagnosis was 0.826 (0.785–0.866), showing moderate diagnostic value similar to mSEPT9.CEA is a glycoprotein mainly present in colorectal cancer tissues and embryonic intestinal mucosa, and its elevation is common in patients with gastrointestinal malignancies, breast cancer, lung cancer, and other malignant tumors ^[28-29]^. CA199 is a gastrointestinal-related carbohydrate antigen that can serve as an auxiliary diagnostic indicator for malignant tumors such as gastric pancreatic cancer and gallbladder cancer. Studies have shown that CA199 increases in upper gastrointestinal tumors such as gastric cancer and duodenal cancer^[30]^. In this study, the levels of CEA and CA199 in the CRC group were both higher than those in the control group (*P*<0.05), with their AUCs (95% CI) for CRC diagnosis being 0.847 (0.808–0.885) and 0.692 (0.641–0.741), indicating moderate and relatively low diagnostic values, respectively.

Inflammation can promote CRC development through mechanisms such as DNA damage, epigenetic alterations, and intestinal epithelial barrier disruption. On the other hand, systemic inflammatory responses accompany both early and advanced cancer, providing new approaches for early cancer identification^[31]^. Compared with single indicators like neutrophils, lymphocytes, platelets, and monocytes, “composite” inflammatory indices such as NLR, PLR, and LMR better reflect the body’s inflammatory level. Studies have shown that these ratio indices are associated with early diagnosis and prognosis in CRC patients^[32-33]^.In this study, NLR and PLR levels were higher in the CRC group than in the control group (*P<*0.001), while LMR levels showed the opposite trend (lower in the CRC group, *P<*0.001), consistent with findings from other researchers^[16]^. In the univariate binary logistic regression model, both NLR and PLR were predictive risk factors for CRC, whereas LMR was a predictive protective factor. When these covariates were included in the multivariate binary logistic regression, only PLR remained a predictive risk factor, while NLR and LMR were excluded from the predictive factors and subsequent diagnostic performance analysis due to statistically insignificant differences (P>0.05). Additionally, the analysis showed that PLR alone had an AUC (95% CI) of 0.670 (0.628–0.713) for CRC diagnosis, indicating relatively low diagnostic value.

RDW, a component of complete blood cell analysis in clinical laboratories, encompasses RDW-CV and RDW-SD. Emerging evidence suggests that RDW in early-stage colorectal cancer (CRC) may reflect the magnitude of tumor-associated inflammatory responses or iron metabolic disorders. Its correlation with clinicopathological features of CRC patients confers potential early prognostic significance^[34]^. Preoperative elevation of RDW has been identified as an independent risk factor for reduced survival in CRC patients ^[35-38]^.This study demonstrated that both RDW-CV and RDW-SD levels were significantly higher in the CRC cohort compared with the control group (*P<*0.05). Notably, RDW-CV was identified as an independent risk factor for CRC development, with a diagnostic efficacy (AUC, 95% CI) of 0.683 (0.640–0.725), comparable to that of PLR, indicating modest diagnostic utility.

This study compared the diagnostic efficacy of plasma mSEPT9 detection with several predictive risk factors for CRC occurrence, evaluating the model’s potential to effectively distinguish between CRC patients and control subjects. It was found that individual plasma mSEPT9 detection exhibited the highest specificity, reaching 0.945 (95% CI 0.926–0.960), while its sensitivity was higher than that of CA199 and RDW-CV. This indicates that plasma mSEPT9 detection holds significant diagnostic value for CRC and may emerge as a promising biomarker for CRC screening. Individual FOBT screening showed the highest sensitivity for CRC but slightly lower specificity, which was lower than that of mSEPT9, CEA, and CA199. Among the clinical efficacy comparisons of individual indicators for CRC diagnosis, CEA demonstrated the highest diagnostic value, with an AUC (95% CI) of 0.847 (0.808– 0.885), confirming that CEA remains an important tool for CRC screening. In this study, individual CA199, RDW-CV, and PLR showed unsatisfactory diagnostic value, with AUC (95% CI) values all below 0.700, suggesting that CA199, RDW-CV, and PLR may not be suitable for independent CRC screening.Furthermore, this study found that combining plasma mSEPT9 with CEA, CA199, FOBT, RDW-CV, or PLR individually improved both AUC and sensitivity compared to single indicators. To the best of our knowledge, no previous studies have reported the combined efficacy of mSEPT9, CEA, CA199, FOBT, RDW-CV, and PLR in CRC diagnosis. Our results showed that the combined detection of these indicators achieved an exceptionally high AUC (95% CI) of 0.939 (0.912–0.966) for CRC diagnosis, with equally remarkable sensitivity and specificity. The AUC and sensitivity of this combined model significantly exceeded those of all single indicators and pairwise combinations, providing surprising diagnostic efficacy in distinguishing between CRC and non-CRC individuals and offering excellent value for non-invasive CRC screening.By integrating measurements of multiple biomarkers to calculate disease probability, such models comprehensively capture the complex multi-layered characteristics of cancer development, outperforming single-analyte detection methods. This study successfully explores the application value of multivariate models in CRC diagnosis, holding promise to further provide clinical practice references for efficient diagnosis and treatment of CRC patients.

Although this study has made some clinically significant discoveries, there are still several limitations. Given that the tumor stages of some clinical CRC patients fall between two stages and cannot be clearly distinguished, this study failed to analyze whether the diagnostic efficacy of plasma mSEPT9 combined with other items would change in CRC patients with different clinical stages, different lesion sites, and different ethnicities. Meanwhile, this study did not evaluate the application value of plasma mSEPT9 combined with other tests in monitoring CRC recurrence before and after treatment, which will be the focus of our subsequent research.

## Conclusions

The combined diagnostic model integrating mSEPT9 with CEA, CA199, FOBT, RDW-CV and PLR demonstrates excellent screening efficacy for CRC, suggesting its potential clinical utility as a comprehensive biomarker panel for early detection.These findings indicate that multimodal biomarker analysis incorporating epigenetic, tumor, hematological and inflammatory markers significantly improves diagnostic accuracy for colorectal cancer compared to individual tests alone.

## Data Availability

All relevant data are within the manuscript and its Supporting Information files.

## Author contributions

**Shuang Yang:** Investigation, Project administration, Writing – original draft. **Yu Qing Wang:** Funding acquisition, Data curation, Methodology. **Jiang Li:** Data curation, Supervision. **Kai Wu Xu:** Project administration, Supervision, Writing – review & editing

## Acknowledgments

The authors extend sincere gratitude to Hunan Provincial People’s Hospital for providing critical clinical data that enabled the exploration of diagnostic markers in this study.

